# Risk of depression, suicidal ideation, suicide and psychosis with hydroxychloroquine treatment for rheumatoid arthritis: a multi-national network cohort study

**DOI:** 10.1101/2020.07.17.20156059

**Authors:** Jennifer C.E. Lane, James Weaver, Kristin Kostka, Talita Duarte-Salles, Maria Tereza F. Abrahao, Heba Alghoul, Osaid Alser, Thamir M Alshammari, Carlos Areia, Patricia Biedermann, Juan M. Banda, Edward Burn, Paula Casajust, Kristina Fišer, Jill Hardin, Laura Hester, George Hripcsak, Benjamin Skov Kaas-Hansen, Sajan Khosla, Spyros Kolovos, Kristine E. Lynch, Rupa Makadia, Paras P. Mehta, Daniel R. Morales, Henry Morgan-Stewart, Mees Mosseveld, Danielle Newby, Fredrik Nyberg, Anna Ostropolets, Rae Woong Park, Albert Prats-Uribe, Gowtham A. Rao, Christian Reich, Peter Rijnbeek, Anthony G. Sena, Azza Shoaibi, Matthew Spotnitz, Vignesh Subbian, Marc A. Suchard, David Vizcaya, Haini Wen, Marcel de Wilde, Junqing Xie, Seng Chan You, Lin Zhang, Simon Lovestone, Patrick Ryan, Daniel Prieto-Alhambra, on behalf of OHDSI-COVID-19 consortium

## Abstract

**Objectives:** Concern has been raised in the rheumatological community regarding recent regulatory warnings that hydroxychloroquine used in the COVID-19 pandemic could cause acute psychiatric events. We aimed to study whether there is risk of incident depression, suicidal ideation, or psychosis associated with hydroxychloroquine as used for rheumatoid arthritis (RA).

**Methods:** New user cohort study using claims and electronic medical records from 10 sources and 3 countries (Germany, UK and US). RA patients aged 18+ and initiating hydroxychloroquine were compared to those initiating sulfasalazine (active comparator) and followed up in the short (30-day) and long term (on treatment). Study outcomes included depression, suicide/suicidal ideation, and hospitalization for psychosis. Propensity score stratification and calibration using negative control outcomes were used to address confounding. Cox models were fitted to estimate database-specific calibrated hazard ratios (HR), with estimates pooled where I^2^<40%.

**Results:** 918,144 and 290,383 users of hydroxychloroquine and sulfasalazine, respectively, were included. No consistent risk of psychiatric events was observed with short-term hydroxychloroquine (compared to sulfasalazine) use, with meta-analytic HRs of 0.96 [0.79-1.16] for depression, 0.94 [0.49-1.77] for suicide/suicidal ideation, and 1.03 [0.66-1.60] for psychosis. No consistent long-term risk was seen, with meta-analytic HRs 0.94 [0.71-1.26] for depression, 0.77 [0.56-1.07] for suicide/suicidal ideation, and 0.99 [0.72-1.35] for psychosis.

**Conclusions:** Hydroxychloroquine as used to treat RA does not appear to increase the risk of depression, suicide/suicidal ideation, or psychosis compared to sulfasalazine. No effects were seen in the short or long term. Use at higher dose or for different indications needs further investigation.

*TRIAL REGISTRATION:* Registered with EU PAS; Reference number EUPAS34497 (http://www.encepp.eu/encepp/viewResource.htm?id=34498). The full study protocol and analysis source code can be found at https://github.com/ohdsi-studies/Covid19EstimationHydroxychloroquine.

*WHAT IS ALREADY KNOWN ON THIS TOPIC:*

- Recent regulatory warnings have raised concerns of potential psychiatric side effects of hydroxychloroquine at the doses used to treat COVID-19, generating concern in the rheumatological community
- Serious psychiatric adverse events such as suicide, acute psychosis, and depressive episodes have been identified by the US Food and Drug Administration (FDA) adverse events reporting system and at case report level

*WHAT THIS STUDY ADDS:*

- This is the largest study on the neuro-psychiatric safety of hydroxychloroquine to date, including >900,000 users treated for their RA in country-level or private health care systems in Germany, the UK, and the US
- We find no association between the use of hydroxychloroquine and the risk of depression, suicide/suicidal ideation, or severe psychosis compared to sulfasalazine

*HOW MIGHT THIS IMPACT ON CLINICAL PRACTICE:*

- Our data shows no association between hydroxychloroquine treatment for RA and risk of depression, suicide or psychosis compared to sulfasalazine. These findings do not support stopping or switching hydroxychloroquine treatment as used for RA due to recent concerns based on COVID-19 treated patients.

## INTRODUCTION

Hydroxychloroquine (HCQ) has received much scientific and public attention during the COVID-19 pandemic as a leading therapeutic and prophylactic target. [1, 2] Commonly used for autoimmune disorders (e.g., systemic lupus erythematosus) and inflammatory arthritis, HCQ was released for emergency use for COVID-19 due to its postulated antiviral efficacy in cellular studies.[3-9] HCQ is currently being used in over 217 registered ongoing clinical trials for the treatment of SARS-Cov-2 as of 12^th^ June 2020.[10, 11] Results to date have been conflicting, with emerging data suggesting a lack of clinical efficacy against COVID-19[12-18]. Potential side effects described in the use of HCQ include neuropsychiatric side effects such as psychosis, depression, and suicidal behaviour.[19-21] Regulatory authorities have received reports of new onset psychiatric symptoms associated with the increased use of high dose HCQ during the pandemic.[22]

New reports of serious side effects associated with HCQ used in COVID-19 are concerning to the rheumatology community, leading to confusion and anxiety for patients who are taking HCQ for autoimmune conditions. We performed a review of the literature to determine what was already known about the potential risks of psychosis, depression, and suicide associated with HCQ use from literature database inception until 14/05/2020 (Supplementary Appendix Section 1). Interrogation of adverse event registers have identified potential associations between HCQ and psychiatric disorders.[11] Case reports and case series describing new onset psychosis, bipolar disorder, seizures and depression associated with HCQ and chloroquine use for rheumatological disorders and malaria prophylaxis can be found as early as 1964.[19, 23-31] No clinical trial or observational study was found that had investigated the incidence of new onset neuropsychiatric symptoms associated with HCQ use.

Considering the wide-scale use of HCQ in rheumatology, we therefore aimed to determine if there is an association between incident HCQ use for rheumatoid arthritis (RA) (the most common indication for the drug) and the onset of acute psychiatric events, including depression, suicide, and psychosis compared to sulfasalazine.

## METHODS

### Study design

A new user cohort, active-comparator design was used, as recommended by methodological guidelines for observational drug safety research.[32] The study protocol is registered in the EU PAS Register as EUPAS34497.[33] Sulfasalazine (SSZ) was used as the active comparator for HCQ,

### Data sources

Electronic health records (EHR) and administrative claims data from the UK and US were used, previously mapped to the Observational Medical Outcomes Partnership (OMOP) common data model (CDM). The study period covered from September 2000 until the latest data available at the time of extraction in each database. Data from 10 data sources were analysed in a federated manner using a distributed network strategy in collaboration with the Observational Health Data Science and Informatics (OHDSI) and European Health Data and Evidence Network (EHDEN) communities. The data used included primary care electronic medical records from the UK (Clinical Practice Research Datalink, CPRD; and IQVIA Medical Research Data, IMRD); specialist ambulatory care electronic health records from Germany (IQVIA Database Analyzer Germany; DAGermany); electronic health records in a sample of US inpatient and outpatient facilities the Optum^®^ de-identified Electronic Health Record dataset (Optum EHR, and IQVIA US Ambulatory EMR;AmbEMR); and US claims data from the IBM MarketScan^®^ Commercial Claims Database (CCAE), Optum^®^ de-identified Clinformatics^®^ Data Mart Database-Date of Death (Clinformatics), IBM MarketScan^®^ Medicare Supplemental Database (MDCR), IBM MarketScan^®^ Multi-State Medicaid Database (MDCD), and IQVIA OpenClaims (OpenClaims). In addition, data were obtained and analysed from electronic primary care data from the Netherlands (IPCI database) and Spain (SIDIAP), and from Japanese claims (JMDC) but none of these analyses were deemed appropriate due to low/no event counts in at least one of the cohorts. A more detailed description of all these data sources is available in Appendix Section 2.

### Follow-up

Participants were followed up from the date of initiation (first dispensing or prescription) of HCQ or sulfasalazine (SSZ) (index date) as described in detail in Appendix Section 3.1. Sulfasalazine was proposed as an active comparator as it shares a similar indication as a second-line conventional synthetic DMARD for RA. Two different follow-up periods were pre-specified to look at short- and long-term effects, respectively. First, a fixed 30-day time window from index date was used to study short-term effects, where follow-up included from day 1 post-index until the earliest of: loss to follow-up/death, outcome of interest, or 30 days from therapy initiation, regardless of compliance/persistence with the study drug/s. Second, in a long-term (on treatment) analysis, follow-up went from day 1 post-index until the earliest of: therapy discontinuation (with a 14-day additional washout), outcome of interest, or loss to follow-up/death. Continued treatment episodes were constructed based on dispensing/prescription records, with a 90-day refill gap allowed to account for stockpiling.

### Participants

All subjects registered in any of the contributing data sources for at least 365 days prior to index date, aged 18 years or older, with a history of RA (as defined by a recorded diagnosis any time before or on the same day as therapy initiation), and starting either HCQ or SSZ during the study period, were included.

Potential participant counts and age-, sex- and calendar year-specific incidence per database were produced for transparency and reviewed to check for data inconsistencies and face validity, and are available for inspection at https://data.ohdsi.org/Covid19CohortEvaluationExposures/, labelled as “New users of hydroxychloroquine with previous rheumatoid arthritis” and “New users of sulfasalazine with previous rheumatoid arthritis”.

### Outcomes and confounders

Code lists for the identification of the study population, for the study exposures and for the relevant outcomes were created by clinicians with experience in the management of RA and by clinical epidemiologists using ATLAS, an open science analytics platform that provides a unified interface for researchers to work within.[34] Exposures and outcomes were reviewed by experts in OMOP vocabulary and in the use of the proposed data sources. A total of three outcomes were analysed: depression, suicide or suicidal ideation, and hospital admission for psychosis. Detailed outcome definitions with links to code lists are fully detailed in Appendix Section 3.2.[35] [36] Cohort counts for each of the outcomes in the entire source database, and age-sex and calendar-time specific incidence rates were explored for each of the contributing databases, and reviewed to check for data inconsistencies and face validity. These are available for inspection at https://data.ohdsi.org/Covid19CohortEvaluationSafetyOutcomes/

A list of negative control outcomes was generated for which there is no biologically plausible or known causal relationship with the use of HCQ or SSZ. These outcomes were identified based on previous literature, clinical knowledge (reviewed by two clinicians), product labels, and spontaneous reports, and confirmed by manual review by two clinicians.*[37]* The full list of codes used to identify negative control outcomes can be found in Appendix Section 4.

### Statistical methods

All analytical source code is available for inspection and reproducibility at https://github.com/ohdsi-studies/Covid19EstimationHydroxychloroquine2. All study diagnostics and the steps described below are available for review at https://data.ohdsi.org/Covid19EstimationHydroxychloroquine2/.

The following steps were followed for each analysis:

#### 1. Propensity score estimation

Propensity score (PS) stratification was used to minimise confounding. All baseline characteristics recorded in the participants’ records/health claims were constructed for inclusion as potential confounders (including demographics, past medical history, procedures and medication prescription within 30 and within 365 days prior to drug initiation) [35]. Covariate construction details are available in Appendix Section 5. Lasso regression models were fitted to estimate propensity scores (PS) as the probability of hydroxychloroquine versus sulfasalazine use based on patient demographics and medical history including previous conditions, procedures, healthcare resource use, and treatments.

The full resulting PS models are available for inspection by clicking on ‘Propensity model’ after selecting a database in the results app.

#### 2. Study diagnostics

Study diagnostics were explored for each database-specific analysis before progressing to outcome modelling, and included checks for power, observed confounding, and potential residual (unobserved) confounding. Only database-outcome analyses that passed all diagnostics below were then conducted and reported, with all others marked as ‘NA’ in the accompanying results app. Positivity and power were assessed by looking at the number of participants in each treatment arm, and the number with the outcome (see the ‘Power’ tab after clicking on a database in the results app). Small cell counts less than five (and resulting estimates) are reported as “<5” to minimise risk of secondary disclosure of data with patient identification. PS overlap was also plotted to visualize positivity issues and can be seen by clicking on ‘Propensity Scores’.

Observed confounding was explored by plotting standardized differences before (X axis) vs after (Y) PS stratification, with standardized differences > 0.1 in the Y axis indicating the presence of unresolved confounding [36]: see by clicking on ‘Covariate balance’ in the results app.

Finally, negative control outcome analyses were assessed to identify systematic error due to residual (unobserved) confounding. The results for these are available in the ‘Systematic error’ tab of the results app. The resulting information was used to calibrate the outcome models using empirical calibration *[37, 38]*.

#### 3. Outcome modelling

Cox proportional hazards models conditioned on the PS strata were fitted to estimate Hazard Ratios (HR) for each psychological outcome in new users of HCQ (vs SSZ). Empirical calibration based on the previously described negative control outcomes was used to minimise any potential residual confounding with calibrated HRs and 95% confidence intervals (CI) estimated[38, 39]. All analyses were conducted for each database separately, with estimates combined in random-effects metaanalysis methods where I^2^ ≤40%.[40] The standard errors of the database-specific estimates were adjusted to incorporate estimate variation across databases, where the across-database variance was estimated by comparing each database-specific result to that of an inverse-variance, fixed-effects meta-analysis. No meta-analysis was conducted where I^2^ for a given drug-outcome pair was >40%.

All analyses were conducted using the CohortMethod package, available at https://ohdsi.github.io/CohortMethod/ and the Cyclops package for PS estimation (https://ohdsi.github.io/Cyclops) [41].

### Data Sharing

Open Science is a guiding principle within OHDSI. As such, we provide unfettered access to all open-source analysis tools employed in this study via https://github.com/OHDSI/, as well as all data and results artefacts that do not include patient-level health information via http://data.ohdsi.org/Covid19EstimationHydroxychloroquine2. Data partners contributing to this study remain custodians of their individual patient-level health information and hold either IRB exemption or approval for participation.

## RESULTS

A total of 918,144 HCQ and 290,383 SSZ users were identified. Participant counts in each data source are provided in Appendix Section 6. Before PS stratification, users of HCQ were (compared to SSZ users) more likely female (for example, 82.0% vs 74.3% in CCAE database) and less likely to have certain comorbidities such as Crohn’s disease (0.6% vs 1.8% in CCAE) or psoriasis (3.0% vs 8.9% in CCAE). Prevalence of systemic lupus erythematous was higher in HCQ users as expected (1.5% vs 0.5% in CCAE), whilst use of systemic glucocorticoids was similar (46.1% vs 47.2% in the previous month in CCAE). The prevalence of depressive disorder was similar in both groups (13.4% vs 13.5% in CCAE) and so was the history of use of antidepressants in the previous year (36.4% vs 36.4% in CCAE). Average baseline dose of HCQ was homogeneous, with >97% in CCAE using an average dose of 420mg daily, and only <3% taking an estimate dose >500 mg. All the observed differences between groups were minimised to an acceptable degree (<0.1 standardised mean differences) after propensity score stratification: in CCAE, the most imbalanced variable was use of glucocorticoids on index date, with prevalence 36.1% vs 35.8%. Detailed baseline characteristics for the two pairs of treatment groups after PS stratification in CCAE are shown in Table 1 as an example, with similar tables and a more extensive list of features provided in Appendix Section 7. Study diagnostics including plots of propensity score distribution, covariate balance, and negative control estimate distributions are provided in Appendix Section 8.

**Table 1.** Baseline characteristics of patients with RA who are new users of hydroxychloroquine (HCQ) vs sulfasalazine (SSZ), before and after PS stratification, in the CCAE database

|  | Before PS stratification |  |  | After PS stratification |  |  |
| --- | --- | --- | --- | --- | --- | --- |
|  | HCQ | SSZ |  | HCQ | SSZ |  |
|  | % | % | Std. diff | % | % | Std. diff |
| ***Socio-demographics*** |  |  |  |  |  |  |
| Age group |  |  |  |  |  |  |
| 15-19 | 0.6 | 0.6 | 0.00 | 0.6 | 0.6 | 0.00 |
| 20-24 | 1.9 | 1.9 | 0.00 | 1.8 | 2.0 | -0.01 |
| 25-29 | 2.6 | 2.6 | 0.00 | 2.5 | 2.8 | -0.01 |
| 30-34 | 4.5 | 4.6 | 0.00 | 4.5 | 4.3 | 0.01 |
| 35-39 | 7.2 | 7.3 | 0.00 | 7.1 | 7.1 | 0.00 |
| 40-44 | 9.8 | 9.5 | 0.01 | 9.7 | 9.5 | 0.00 |
| 45-49 | 13.7 | 12.9 | 0.02 | 13.6 | 13.5 | 0.00 |
| 50-54 | 18.2 | 18.2 | 0.00 | 18.2 | 18.1 | 0.00 |
| 55-59 | 20.6 | 21.0 | -0.01 | 20.8 | 20.8 | 0.00 |
| 60-64 | 19.0 | 19.7 | -0.02 | 19.4 | 19.8 | -0.01 |
| 65-69 | 1.8 | 1.7 | 0.01 | 1.8 | 1.6 | 0.01 |
| Gender: female | 82.0 | 74.3 | 0.19 | 80.1 | 79.7 | 0.01 |
| ***Medical history*** |  |  |  |  |  |  |
| Acute respiratory disease | 35.5 | 34.3 | 0.03 | 35.1 | 34.7 | 0.01 |
| Chronic liver disease | 3.2 | 3.2 | 0.00 | 3.2 | 3.4 | -0.01 |
| Chronic obstructive lung disease | 4.2 | 4.5 | -0.01 | 4.3 | 4.5 | -0.01 |
| Crohn's disease | 0.6 | 1.8 | -0.12 | 0.7 | 1.1 | -0.04 |
| Depressive disorder | 13.4 | 13.5 | 0.00 | 13.2 | 13.4 | -0.01 |
| Diabetes mellitus | 13.5 | 13.4 | 0.00 | 13.6 | 13.7 | 0.00 |
| Hypertensive disorder | 34.7 | 34.9 | 0.00 | 34.7 | 35.0 | -0.01 |
| Obesity | 9.3 | 9.1 | 0.00 | 9.2 | 9.4 | -0.01 |
| Psoriasis | 3.0 | 8.9 | -0.25 | 3.8 | 5.2 | -0.07 |
| Renal impairment | 3.1 | 2.8 | 0.02 | 3.0 | 2.8 | 0.01 |
| Systemic Lupus Erythematosus | 1.5 | 0.5 | 0.10 | 1.3 | 0.9 | 0.03 |
| Schizophrenia | 0.1 | 0.1 | -0.01 | 0.1 | 0.1 | -0.01 |
| Ulcerative colitis | 0.6 | 1.9 | -0.12 | 0.7 | 1.0 | -0.04 |
| **Medication use** |  |  |  |  |  |  |
| Agents acting on the renin-angiotensin system | 24.3 | 24.9 | -0.01 | 24.5 | 24.7 | 0.00 |
| Antidepressants | 36.4 | 36.4 | 0.00 | 36.3 | 36.5 | 0.00 |
| Antiepileptics | 20.3 | 21.0 | -0.02 | 20.4 | 20.2 | 0.00 |
| Antiinflammatory and antirheumatic products | 55.3 | 57.3 | -0.04 | 55.8 | 56.7 | -0.02 |
| Antipsoriatics | 0.7 | 1.3 | -0.06 | 0.7 | 1.0 | -0.03 |
| Antithrombotic agents | 7.4 | 7.3 | 0.01 | 7.4 | 7.3 | 0.00 |
| Immunosuppressants | 39.6 | 53.1 | -0.27 | 43.4 | 43.6 | 0.00 |
| Opioids | 38.5 | 40.8 | -0.05 | 39.0 | 39.3 | -0.01 |
| Psycholeptics | 33.6 | 33.7 | 0.00 | 33.4 | 33.3 | 0.00 |
| HCQ=hydroxychloroquine; SSZ=sulfasalazine |  |  |  |  |  |  |

Database-specific and overall counts and rates of the three study outcomes in the short- (30-day) and long-term (‘on treatment’) analyses are reported in detail in Table *2*. Depression was the most common of the three study outcomes, with rates in the ‘on treatment’ analysis ranging from 1.99/1,000 person-years amongst HCQ users in CPRD to 17.74/1,000 amongst HCQ users in AmbEMR. Suicide/suicidal ideation was the least common outcome, with rates ranging from 0.32/1,000 (HCQ users in AmbEMR and SSZ users in IMRD) to 14.08/1,000 in SSZ users in MDCD. Database-specific counts and incidence rates (IR) for all three outcomes stratified by drug use are detailed in full in Appendix Section 9.

**Table 2.** Patient counts, event counts and incidence rates (IR) (/1,000 person years) of key events according to drug use

| 30-day follow up |  |  |  |  |  |  |  | On-treatment follow up |  |  |  |  |  |
| --- | --- | --- | --- | --- | --- | --- | --- | --- | --- | --- | --- | --- | --- |
|  |  | Patients |  | Events |  | IR<br>(/1,000<br>py |  | Patients |  | Events |  | IR<br>(/1,000<br>py |  |
| Outcome | Database | T | C | T | C | T | C | T | C | T | C | T | C |
| Depression | AmbEMR | 55,793 | 15,092 | 155 | 29 | 33.91 | 23.44 | 55,793 | 15,092 | 320 | 80 | 17.74 | 14.34 |
|  | CCAE | 66,440 | 22,449 | 79 | 28 | 14.64 | 15.36 | 66,440 | 22,449 | 557 | 137 | 8.54 | 9.40 |
|  | Clinformatics | 51,676 | 16,812 | 84 | 41 | 20.05 | 30.09 | 51,676 | 16,812 | 657 | 178 | 12.43 | 15.00 |
|  | CPRD | 9,160 | 11,348 | <5 | 8 | <6.67 | 8.60 | 9,160 | 11,348 | 36 | 94 | 1.99 | 3.60 |
|  | DAGermany | 3,937 | 5,109 | <5 | 12 | <15.48 | 28.63 | 3,937 | 5,109 | 40 | 70 | 15.47 | 19.66 |
|  | IMRD | 8,844 | 8,456 | <5 | 6 | <6.91 | 8.67 | 8,844 | 8,456 | 38 | 51 | 2.20 | 2.72 |
|  | MDCD | 7,950 | 2,286 | 14 | 6 | 21.61 | 32.29 | 7,950 | 2,286 | 90 | 13 | 15.81 | 10.12 |
|  | MDCR | 15,735 | 5,275 | 13 | 6 | 10.14 | 13.98 | 15,735 | 5,275 | 97 | 38 | 5.37 | 9.27 |
|  | OpenClaims | 620,081 | 183,312 | 654 | 161 | 12.85 | 10.70 | 620,081 | 183,312 | 4,810 | 957 | 5.59 | 5.58 |
|  | OptumEHR | 78,528 | 20,244 | 321 | 66 | 50.56 | 40.30 | NA | NA | NA | NA | NA | NA |
|  | Meta-analysis | 918,144 | 290,383 | <1,335 | 363 | <17.77 | 15.28 | 839,616 | 270,139 | 6,645 | 1,618 | 6.28 | 6.29 |
| Suicide and suicidal ideation | AmbEMR | NA | NA | NA | NA | NA | NA | 57,660 | 15,357 | 6 | <5 | 0.32 | <0.88 |
|  | CCAE | 66,533 | 22,471 | 12 | <5 | 2.22 | <2.74 | 66,533 | 22,471 | 81 | 28 | 1.23 | 1.91 |
|  | Clinformatics | 51,807 | 16,843 | 12 | <5 | 2.85 | <3.66 | 51,807 | 16,843 | 97 | 30 | 1.80 | 2.50 |
|  | CPRD | 9,167 | 11,358 | <5 | <5 | <6.66 | <5.37 | 9,167 | 11,358 | 7 | 9 | 0.39 | 0.34 |
|  | IMRD | 8,852 | 8,460 | <5 | <5 | <6.91 | <7.22 | 8,852 | 8,460 | 8 | 6 | 0.46 | 0.32 |
|  | MDCD | 7,980 | 2,296 | <5 | <5 | <7.68 | <26.78 | 7,980 | 2,296 | 56 | 18 | 9.71 | 14.08 |
|  | MDCR | NA | NA | NA | NA | NA | NA | 15,752 | 5,278 | 15 | 6 | 0.83 | 1.45 |
|  | OpenClaims | 621,067 | 183,550 | 34 | 8 | 0.67 | 0.53 | 621,067 | 183,550 | 321 | 89 | 0.37 | 0.52 |
|  | OptumEHR | 79,903 | 20,480 | 18 | 8 | 2.78 | 4.82 | NA | NA | NA | NA | NA | NA |
|  | Meta-analysis | 845,309 | 265,458 | <91 | <41 | <1.31 | <1.89 | 838,818 | 265,613 | 591 | <191 | 0.55 | <0.75 |
| Hospitalization for psychosis | OpenClaims | 620,964 | 183,527 | 95 | 27 | 1.86 | 1.79 | 620,964 | 183,527 | 1,108 | 221 | 1.28 | 1.28 |
|  | OptumEHR | 79,994 | 20,508 | <5 | <5 | <0.77 | <3.01 | NA | NA | NA | NA | NA | NA |
|  | Meta-analysis | 700,958 | 204,035 | <100 | <32 | <1.74 | <1.91 | NA | NA | NA | NA | NA | NA |

T=target therapy; C=comparator therapy; IR=incidence rate; py=person-years at risk; NA=non-applicable (not reported because of failed diagnostics or on-treatment follow-up unavailable); HCQ=hydroxychloroquine; SSZ=sulfasalazine; AmbEMR=IQVIA Ambulatory EMR; CCAE=IBM Commercial Database; Clinformatics=Optum de-identified Clinformatics Data Mart Database; CPRD=Clinical Practice Research Datalink; DAGermany=IQVIA Disease Analyzer Germany; IMRD=IQVIA UK Integrated Medical Record Data; MDCCD=IBM IBM Multi-state Medicaid; MDCR=IBM Medicare Supplemental Database; OpenClaims=IQVIA Open Claims; OptumEHR=Optum de-identified Electronic Health Record dataset

9 datasets passed cohort diagnostics and contained sufficiently robust data for inclusion into the short term analyses for depression; 6 passed for suicide and 2 passed for psychosis. A small imbalance with the incidence of a past medical history of SLE was seen in MDCD and with cutaneous lupus in DAGermany. As a result, we excluded both from the psychosis outcome but not for depression as we did not consider this was a confounder. Short-term (30-day) analyses showed no consistent association between HCQ use and the risk of depression, with database-specific HRs ranging from 0.21 [95%CI 0.03-1.25] in CPRD to 1.28 [0.85-1.95] in AmbEMR, and a meta-analytic HR of 0.96 [0.79-1.16] (See Figure 1, top). On-treatment analyses showed similar findings, with database-specific HRs from 0.62 [0.40-0.97] in DAGermany to 1.29 [0.69-2.39] in MDCD, and a meta-analytic HR of 0.94 [0.71-1.26] (Figure 1, bottom plot). Note only databases passing diagnostics are included within the plot and meta-analysis.

**Figure 1.**
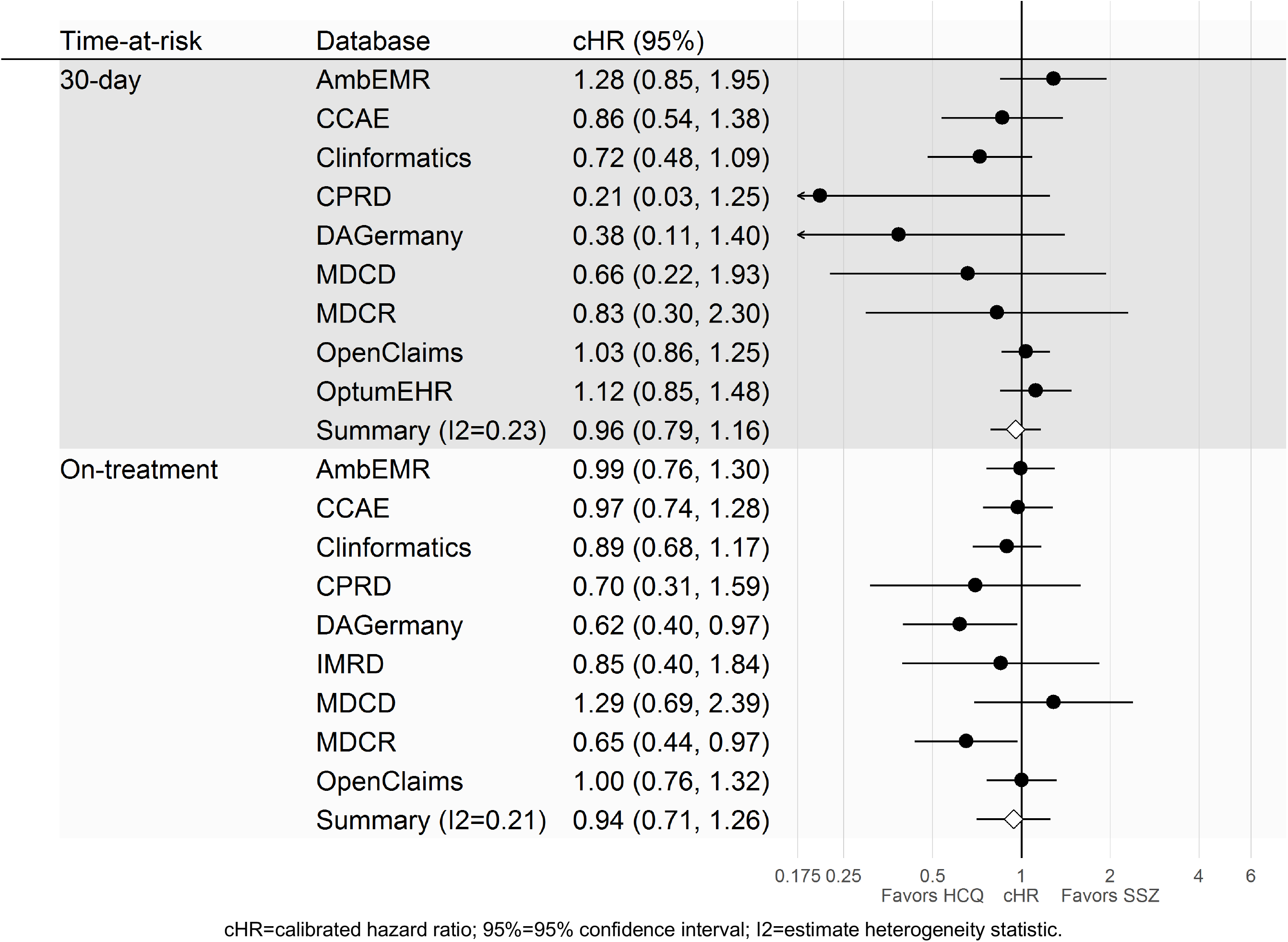
Forest plot of the association between short-(top) and long-term (bottom) use of HCQ (vs SSZ) and risk of depression, by database and in meta-analysis.

Similarly, no association was seen between the use of HCQ and the risk of suicidal ideation or suicide. In the short-term, HRs ranged from 0.27 [0.06-1.29] in MDCD to 10.46 [0.51-216.29] in CPRD, with meta-analytic HR of 0.94 [0.49-1.77] (Figure 2, top). Long-term effects were similar, with HRs ranging between 0.55 [0.20-1.49] in MDCR and 2.36 [0.21-26.87] in AmbEMR, and meta-analytic HR of 0.77 [0.56-1.07] (Figure 2, bottom).

**Figure 2.**
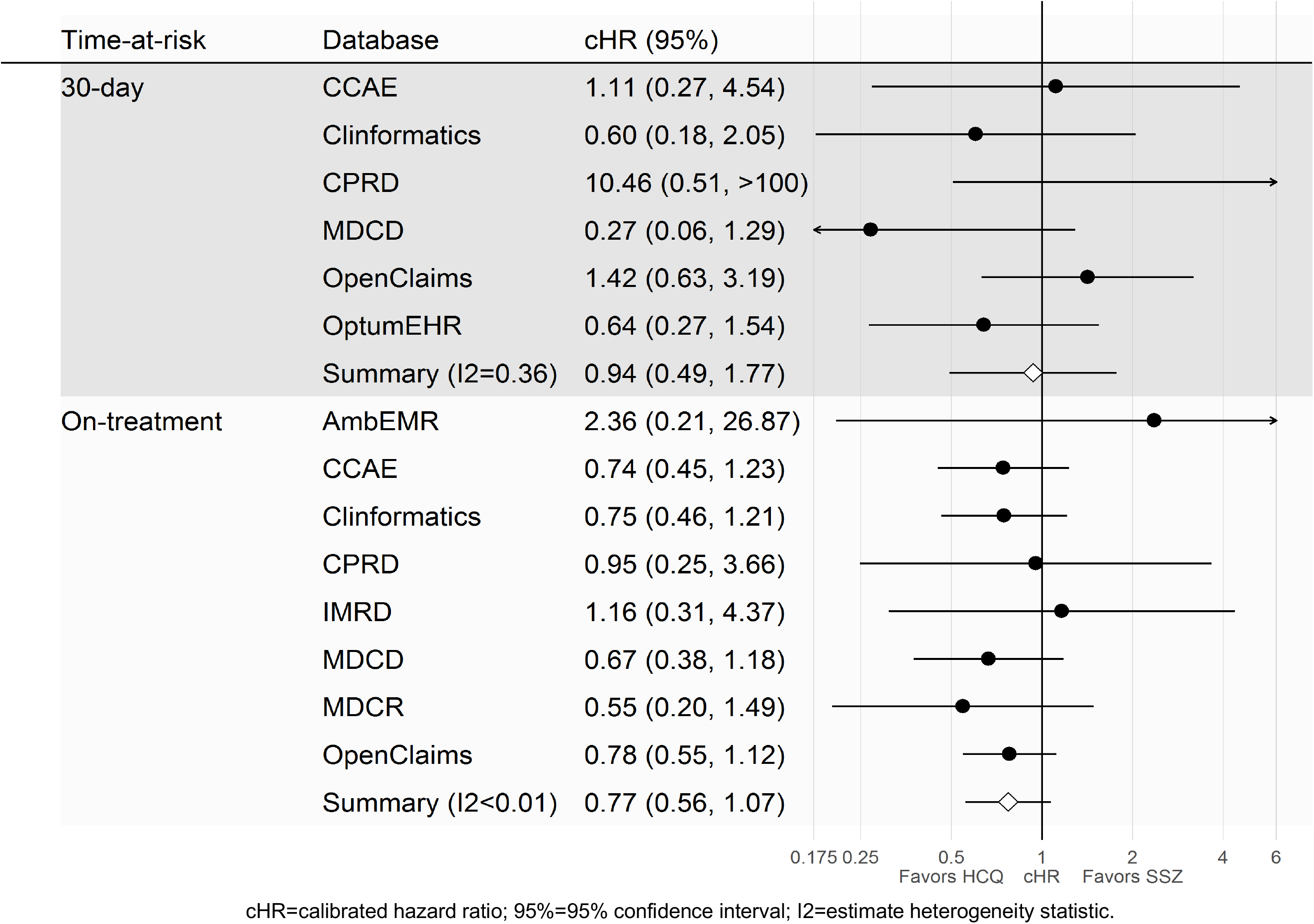
Forest plot of the association between short-(top) and long-term (bottom) use HCQ (vs SSZ) and risk of suicidal ideation or suicide, by database and in meta-analysis.

Finally, no association was seen between the use of HCQ (compared to SSZ) and the risk of acute psychosis. Short-term analyses showed database-specific HRs of 0.44 [0.05-3.49] in OptumEHR and 1.01 [0.65-1.58] in OpenClaims, with a meta-analytic estimated HR of 1.03 [0.66-1.60]. Only OpenClaims contributed to the ‘on treatment’ analysis of this event, with an estimated HR of 0.98 [0.73-1.33].

## DISCUSSION

### Principal findings

This large observational study shows that in routine healthcare treatment of RA, there is no association with the use of HCQ with acute psychosis, depression, or suicide as compared to SSZ. These results are seen both in the short-term and long-term risk analyses. Whilst an excess of psychiatric events have been reported during the COVID pandemic in those prescribed HCQ, this risk does not appear to be associated with HCQ prescribed in RA compared to those prescribed SSZ. This study uses data from three countries, with a variety of healthcare systems and modes of routine healthcare data included, enabling the study to produce more generalisable results.

### Comparison with other studies

The bulk of the evidence prior to this study consisted of isolated case reports and case series, making it difficult to draw demographic comparisons with previous work. Sato *et al*. reported that neuropsychiatric adverse events found in the FDA adverse event reporting system associated with chloroquine use were predominantly in females in the sixth decade of life.[20]Increase in reporting of acute psychiatric disease during the COVID-19 pandemic may be multifactorial, with an increase in external stressors such as social isolation, financial uncertainty, and increased misuse of drugs and alcohol.[42-44] Considering that we find no association for HCQ use compared to SSZ with acute psychiatric outcomes in the RA population, evidence points towards external stressors being more likely involved in the aetiology of psychiatric events seen during this pandemic.

### Strengths and weaknesses of the study

This study is based on new users of HCQ for RA and therefore, the results of this study are most directly relevant to the risk of neuropsychiatric side effects seen in the rheumatological population. The regulatory warnings of possibly increased acute psychiatric events associated with HCQ warrant investigation in all available datasets to prevent harm in both rheumatological patients and those taking for emergency use, especially as very few clinical trials include acute psychiatric outcomes. Whilst the general population presenting with COVID-19 may differ from those with RA, within the context of emergency authorisation or off label use of HCQ, all available evidence must be taken into account when considering the risks associated.

Several considerations must be taken into account when interpreting these results.

Firstly, the doses used to treat RA are lower than those suggested in current clinical trials for the treatment of SARS-CoV2, and therefore adverse events seen in the treatment and prophylaxis of COVID-19 may be greater if dose dependent, as is the case with cardiac adverse effects.[45, 46] Secondly, this study could be affected by outcome misclassification. Only acute psychiatric events presenting to medical services will be captured, and this is especially important for the outcome of suicide. Suicide may not be fully recorded if patients do not reach medical care or cause-of-death information is not linked to the datasource, and therefore the true incidence of suicide may be under-recorded.[47] Similarly, this study only focused on acute psychosis and depression severe enough to be identified in medical consultation in patients with no history of either condition. Whilst we generated phenotypes that underwent full cohort diagnostics, and phenotypes were constructed using a multidisciplinary team of clinicians and bioinformaticians to ensure face validity, it should be noted that no formal validation was undertaken. We took all reasonable steps to ensure the validity of the phenotypes, whilst considering the risk-benefit tradeoff of what could be undertaken within the time frame used to respond to the serious questions raised by regulatory bodies following the HCQ use in COVID-19.

This study can highlight the association for patients without a prior history of psychosis or depression, but cannot inform of the risk of acute deterioration after beginning HCQ treatment for those already known to psychiatric services.

Thirdly, depression and hallucinations are listed as potential undesirable effects of sulfasalazine treatment, which may underestimate the true risk, if any, from HCQ.[48] However, the frequency of depression (described as changes in affect in the summary of product characteristics for HCQ) is reported to be common (≥1/100 to < 1/10) whilst for sulfasalazine depression is listed as being uncommon (≥1/1000 to < 1/100). Therefore, it is potentially reassuring for patients that we observed no difference compared to sulfasalazine for which there is a paucity of published evidence suggesting causailty.[49]

Propensity score stratification and matching, as well as a comprehensive examination of potential sources of systematic error, were undertaken prior to blinding of results to identify and reduce the risk of confounding. Baseline characteristics after PS stratification were adequately balanced; of note, the incidence of systemic lupus erythematosus (SLE) was balanced between treatment groups. Identifying the balance of SLE between treatment groups was undertaken prior to unblinding due to the potential neuropsychiatric sequelae of the condition aside from the potential side effects of pharmacological treatment. This study could also be limited by the fact that patients may overlap and exist in more than one dataset within the US. The meta-analysis assumes populations to be independent, and therefore the obtained estimates may slightly underestimate variance.

### Future research

For rheumatological disorders, future work could expand into investigating the occurrence of acute psychiatric events in patients in SLE. This would enable greater understanding of whether neuropsychiatric conditions are related to disease activity or due to pharmacological treatment. Similarly, in the emergency use of HCQ in COVID-19, there is already concern about the potential heightened risk of acute psychiatric disorder due to elevated number of psychosocial stressors present during a pandemic and high dose use.[50] Future work should consider including acute psychiatric outcomes in order to differentiate between psychiatric conditions generated by the impact of a global pandemic compared to iatrogenic events due to pharmaceutical therapies used.

### Meaning of the Study

Exponential growth in research into the best treatment of SARS-CoV2 infection is generating rapidly evolving evidence for the relative efficacy of pharmaceutical agents. For the rheumatological community, media attention previously surrounded HCQ as a strong forerunner of COVID-19 prophylaxis and treatment. The results of the RECOVERY trial identifying dexamethasone reduced mortality in intensive care patients has now overtaken HCQ as the leading rheumatological drug for the pandemic, but the concerns regarding HCQ safety remain for those who take the drug for conventional indications.[17, 51] Cardiovascular safety, and reports that it might lack efficacy for both treatment and prophylaxis, have halted major HCQ clinical trials.[45, 52-55] The identification of acute psychiatric events associated with HCQ use has raised the need to clarify the risk within general rheumatological use. Our study identifies no increased risk in RA patients when compared with sulfasalazine, and provides evidence to users and clinicians alike that the reports presented during the pandemic are likely to be related to further causes aside from HCQ.

## Data Availability

This study was conducted as a distributed database network analysis. To protect patient privacy, all patient-level data were maintained securely behind institutional firewalls. Analysis code was downloaded and executed by each participating data partner, which generated only aggregate summary statistics (cohort counts, model coefficients) which were then centralized and synthesized in preparation of this manuscript. All aggregate summary statistics produced have been made publicly available at: https://data.ohdsi.org/Covid19EstimationHydroxychloroquine2/.

## FOOTNOTES

## Acknowledgements

Catherine Hartley and Eli Harriss of Bodleian Health Care Libraries, University of Oxford; Runsheng Wang, Joel Swerdel, Zeshan Ghory, Liliana Ciobanu, Michael Kallfelz, Nigel Hughes and Martijn Schuemie, Mitchell M. Conover, Aedin C. Culhane Scott L. DuVall, Dmitry Dymshyts, Seamus Kent, Christophe G. Lambert, Johan van der Lei, Andrea V. Margulis, Michael E. Matheny, Lisa Schilling, Sarah Seager, and Oleg Zhuk. Finally, we acknowledge the tremendous work and dedication of the 350 participants from 30 nations in the March 2020 OHDSI COVID-19 Virtual Study-a-thon (https://www.ohdsi.org/covid-19-updates/), without whom this study could not have been realized.

## Public and patient involvement

No patients were directly involved in setting the research question, nor in design, conduct or interpretation of the study.

## Ethical Approval

All data partners received IRB approval or waiver in accordance to their institutional governance guidelines.

| **Database** | **Statement** |
| --- | --- |
| AmbEMR | This is a retrospective database study on de-identified data and is deemed not human subject research. Approval is provided for OHDSI community studies. |
| CCAE | New England Institutional Review Board (IRB) and was determined to be exempt from broad IRB approval, as this research project did not involve human subject research. |
| CPRD | Approval for CPRD was provided by the Independent Scientific Advisory Committee (ISAC).<br>This study is based in part on data from the Clinical Practice Research Datalink obtained under licence from the UK Medicines and Healthcare products Regulatory Agency. The data is provided by patients and collected by the NHS as part of their care and support. The interpretation and |
|  | conclusions contained in this study are those of the author/s alone. The protocol for this study ( 20_059R) was approved by the Independent Scientific Advisory Committee (ISAC). |
| DA Germany | This is a retrospective database study on de-identified data and is deemed not human subject research. Approval is provided for OHDSI community studies. |
| IMRD | The present study is filed and under review for Scientific Review Committee for institutional adjudication. Due to the public health imperative of information related to these data, approval is provided for this publication. |
| IPCI | The present study was approved by the Scientific and Ethical Advisory Board of the IPCI project (project number: 4/2020). |
| JMDC | New England Institutional Review Board (IRB) and was determined to be exempt from broad IRB approval, as this research project did not involve human subject research. |
| MDCD | New England Institutional Review Board (IRB) and was determined to be exempt from broad IRB approval, as this research project did not involve human subject research. |
| MDCD | New England Institutional Review Board (IRB) and was determined to be exempt from broad IRB approval, as this research project did not involve human subject research. |
| Open Claims | This is a retrospective database study on de-identified data and is deemed not human subject research. Approval is provided for OHDSI community studies. |
| Clinformatics | New England Institutional Review Board (IRB) and was determined to be exempt from broad IRB approval, as this research project did not involve human subject research. |
| Optum EHR | New England Institutional Review Board (IRB) and was determined to be exempt from broad IRB approval, as this research project did not involve human subject research. |

## REFERENCES

1. Yao X, Ye F, Zhang M, Cui C, Huang B, Niu P, et al. In Vitro Antiviral Activity and Projection of Optimized Dosing Design of Hydroxychloroquine for the Treatment of Severe Acute Respiratory Syndrome Coronavirus 2 (SARS-CoV-2). Clin Infect Dis. 2020.

2. Ferner RE, Aronson JK. Chloroquine and hydroxychloroquine in covid-19. Bmj. 2020;369:m1432.

3. Sepriano A, Kerschbaumer A, Smolen JS, van der Heijde D, Dougados M, van Vollenhoven R, et al. Safety of synthetic and biological DMARDs: a systematic literature review informing the 2019 update of the EULAR recommendations for the management of rheumatoid arthritis. Ann Rheum Dis. 2020.

4. FDA. Request for Emergency Use Authorization For Use of Chloroquine Phosphate or Hydroxychloroquine Sulfate Supplied From the Strategic National Stockpile for Treatment of 2019 Coronavirus Disease. In: Administration UFaD, editor. Bethseda, USA 2020.

5. EMA. COVID-19: chloroquine and hydroxychloroquine only to be used in clinical trials or emergency use programmes. Amsterdam 2020.

6. Colson P, Rolain J-M, Lagier J-C, Brouqui P, Raoult D. Chloroquine and hydroxychloroquine as available weapons to fight COVID-19. International journal of antimicrobial agents. 2020:105932-.

7. Liu J, Cao R, Xu M, Wang X, Zhang H, Hu H, et al. Hydroxychloroquine, a less toxic derivative of chloroquine, is effective in inhibiting SARS-CoV-2 infection in vitro. Cell Discov. 2020;6:16.

8. Vigerust DJ, Shepherd VL. Virus glycosylation: role in virulence and immune interactions. Trends in Microbiology. 2007;15(5):211-8.

9. Wang M, Cao R, Zhang L, Yang X, Liu J, Xu M, et al. Remdesivir and chloroquine effectively inhibit the recently emerged novel coronavirus (2019-nCoV) in vitro. Cell Res. 2020;30(3):269-71.

10. Gao J, Hu S. Update on use of chloroquine/hydroxychloroquine to treat coronavirus disease 2019 (COVID-19). Biosci Trends. 2020.

11. Randomised Evaluation of COVID-19 Therapy (RECOVERY) [Internet]. 2020. Available from: https://clinicaltrials.gov/ct2/show/NCT04381936 [last accessed 12.06.2020].

12. Boulware DR, Pullen MF, Bangdiwala AS, Pastick KA, Lofgren SM, Okafor EC, et al. A Randomized Trial of Hydroxychloroquine as Postexposure Prophylaxis for Covid-19. N Engl J Med. 2020.

13. Molina JM, Delaugerre C, Goff JL, Mela-Lima B, Ponscarme D, Goldwirt L, et al. No Evidence of Rapid Antiviral Clearance or Clinical Benefit with the Combination of Hydroxychloroquine and Azithromycin in Patients with Severe COVID-19 Infection. Med Mal Infect. 2020.

14. Gautret P, Lagier JC, Parola P, Hoang VT, Meddeb L, Mailhe M, et al. Hydroxychloroquine and azithromycin as a treatment of COVID-19: results of an open-label non-randomized clinical trial. Int J Antimicrob Agents. 2020:105949.

15. Mahévas M, Tran VT, Roumier M, Chabrol A, Paule R, Guillaud C, et al. Clinical efficacy of hydroxychloroquine in patients with covid-19 pneumonia who require oxygen: observational comparative study using routine care data. BMJ. 2020;369:m1844.

16. Gao J, Tian Z, Yang X. Breakthrough: Chloroquine phosphate has shown apparent efficacy in treatment of COVID-19 associated pneumonia in clinical studies. Biosci Trends. 2020;14(1):72-3.

17. No clinical benefit from the use of hydroxychloroquine in hospitalised patients with COVID-19 [press release]. 2020.

18. WHO. “Solidarity” clinical trial for COVID-19 treatments 2020 [Available from: https://www.who.int/emergencies/diseases/novel-coronavirus-2019/global-research-on-novel-coronavirus-2019-ncov/solidarity-clinical-trial-for-covid-19-treatments [last accessed 12.06.2020].

19. Mascolo A, Berrino PM, Capuano A, Berrino L, Gareri P, Castagna A, et al. Neuropsychiatric clinical manifestations in elderly patients treated with hydroxychloroquine: a review article. Inflammopharmacology. 2018;26(5):1141-9.

20. Sato K, Mano T, Iwata A, Toda T. Neuropsychiatric adverse events of chloroquine: a real-world pharmacovigilance study using the FDA Adverse Event Reporting System (FAERS) database. Biosci Trends. 2020.

21. EMA. COVID-19: reminder of the risks of chloroquine and hydroxychloroquine. 2020.

22. AEMPS. Cloroquina/Hidroxicloroquina: precauciones y vigilancia de posibles reacciones adversas en pacientes con COVID-19-Trastornos neuropsiquiatricos. 2020.

23. Gurbuz-Ozgur B, Aslan-Kunt D, Sevincok L. Psychotic symptoms related to hydroxychloroquine. Klinik Psikofarmakoloji Bulteni. 2014;24.

24. Manzo C, Gareri P, Castagna A. Psychomotor Agitation Following Treatment with Hydroxychloroquine. Drug Saf Case Rep. 2017;4(1):6.

25. Sapp OL, 3rd. TOXIC PSYCHOSIS DUE TO QUINARCRINE AND CHLOROQUINE. JAMA. 1964;187:373-5.

26. Bogaczewicz A, Sobow T, Bogaczewicz J, Bienkowski P, Kowalski J, Wozniacka A. Chloroquine-induced subacute paranoid-like disorder as a complication of dermatological treatment. Int J Dermatol. 2016;55(12):1378-80.

27. Pinho De Oliveira Ribeiro N, Rafael De Mello Schier A, Ornelas AC, Nardi AE, Silva AC, Pinho De Oliveira CM. Anxiety, depression and suicidal ideation in patients with rheumatoid arthritis in use of methotrexate, hydroxychloroquine, leflunomide and biological drugs. Comprehensive Psychiatry. 2013;54(8):1185-9.

28. Fish DR, Espir ML. Convulsions associated with prophylactic antimalarial drugs: implications for people with epilepsy. BMJ. 1988;297(6647):526-7.

29. Ward WQ, Walter-Ryan WG, Shehi GM. Toxic psychosis: a complication of antimalarial therapy. J Am Acad Dermatol. 1985;12(5 Pt 1):863-5.

30. Gonzalez-Nieto JA, Costa-Juan E. Psychiatric symptoms induced by hydroxychloroquine. Lupus. 2015;24(3):339-40.

31. Hsu W, Chiu N, Huang S. Hydroxychloroquine-induced acute psychosis in a systemic lupus erythematosus female. Acta Neuropsychiatr. 2011;23(6):318-9.

32. EMA. ENCePP Guide on Methodological Standards in Pharmacoepidemiology. In: ENCePP, editor. 2020.

33. EMA. EU PAS registration: Hydroxychloroquine safety and potential efficacy as an antiviral prophylaxis in light of potential wide-spread use in COVID-19: a multinational, large-scale network cohort and self-controlled case series study. In: ENCEPP, editor. 2020.

34. OHDSI. ATLAS open science analytics platform 2020 [Available from: https://atlas.ohdsi.org/#/home [last accessed 25.06.2020].

35. OHDSI. The Book of OHDSI2020.

36. Suchard MA, Schuemie MJ, Krumholz HM, You SC, Chen R, Pratt N, et al. Comprehensive comparative effectiveness and safety of first-line antihypertensive drug classes: a systematic, multinational, large-scale analysis. Lancet. 2019;394(10211):1816-26.

37. Voss EA, Boyce RD, Ryan PB, van der Lei J, Rijnbeek PR, Schuemie MJ. Accuracy of an automated knowledge base for identifying drug adverse reactions. J Biomed Inform. 2017;66:72-81.

38. Schuemie MJ, Hripcsak G, Ryan PB, Madigan D, Suchard MA. Robust empirical calibration of p-values using observational data. Stat Med. 2016;35(22):3883-8.

39. Schuemie MJ, Ryan PB, DuMouchel W, Suchard MA, Madigan D. Interpreting observational studies: why empirical calibration is needed to correct p-values. Stat Med. 2014;33(2):209-18.

40. DerSimonian R, Laird N. Meta-analysis in clinical trials. Control Clin Trials. 1986;7(3):177-88.

41. Suchard MA, Simpson SE, Zorych I, Ryan P, Madigan D. Massive parallelization of serial inference algorithms for a complex generalized linear model. ACM Trans Model Comput Simul. 2013;23(1).

42. Yao H, Chen JH, Xu YF. Patients with mental health disorders in the COVID-19 epidemic. Lancet Psychiatry. 2020;7(4):e21.

43. Stuckler D, Basu S, Suhrcke M, Coutts A, McKee M. The public health effect of economic crises and alternative policy responses in Europe: an empirical analysis. Lancet. 2009;374(9686):315-23.

44. O’Connor RC, Kirtley OJ. The integrated motivational-volitional model of suicidal behaviour. Philos Trans R Soc Lond B Biol Sci. 2018;373(1754).

45. Mercuro NJ, Yen CF, Shim DJ, Maher TR, McCoy CM, Zimetbaum PJ, et al. Risk of QT Interval Prolongation Associated With Use of Hydroxychloroquine With or Without Concomitant Azithromycin Among Hospitalized Patients Testing Positive for Coronavirus Disease 2019 (COVID-19). JAMA Cardiol. 2020.

46. ACC. Ventricular Arrhythmia Risk due to hydroxychloroquine-azithromycin treatment for COVID-19 2020 [Available from: https://www.acc.org/latest-in-cardiology/articles/2020/03/27/14/00/ventricular-arrhythmia-risk-due-to-hydroxychloroquine-azithromycin-treatment-for-covid-19 [last accessed 25.06.2020].

47. Thomas KH, Davies N, Metcalfe C, Windmeijer F, Martin RM, Gunnell D. Validation of suicide and self-harm records in the Clinical Practice Research Datalink. Br J Clin Pharmacol. 2013;76(1):145-57.

48. EMC. Salazopyrin 2020 [Available from: https://www.medicines.org.uk/emc/product/3838/smpc [last accessed 25.06.2020].

49. Jajić Z, Jajić I. [Acute psychoses in patients with psoriatic arthritis during treatment with sulfasalazine]. Reumatizam. 1998;46(1):43-4.

50. Gunnell D, Appleby L, Arensman E, Hawton K, John A, Kapur N, et al. Suicide risk and prevention during the COVID-19 pandemic. Lancet Psychiatry. 2020;7(6):468-71.

51. Horby P, Lim WS, Emberson J, Mafham M, Bell J, Linsell L, et al. Effect of Dexamethasone in Hospitalized Patients with COVID-19: Preliminary Report. medRxiv. 2020:2020.06.22.20137273.

52. Luo MH, Q. Guirong, X. Wu, F. Wu, B. Xu, T. Data Mining and Safety Analysis of Drugs for Novel Coronavirus Pneumonia Treatment based on FAERS: Chloroquine Phosphate Herald of Medicine (Yi Yao Dao Bao). 2020;2020-02-29 online first:1-14.

53. Roden DM, Harrington RA, Poppas A, Russo AM. Considerations for Drug Interactions on QTc in Exploratory COVID-19 (Coronavirus Disease 2019) Treatment. Circulation. 2020.

54. Lane JCE, Weaver J, Kostka K, Duarte-Salles T, Abrahao MTF, Alghoul H, et al. Safety of hydroxychloroquine, alone and in combination with azithromycin, in light of rapid wide-spread use for COVID-19: a multinational, network cohort and self-controlled case series study. medRxiv. 2020:2020.04.08.20054551.

55. NIH. NIH halts clinical trial of hydroxychloroquine 2020 [Available from: https://www.nih.gov/news-events/news-releases/nih-halts-clinical-trial-hydroxychloroquine [last accessed 25.06.2020].

